# Genetic and functional characterization of inherited complex chromosomal rearrangements in a family with multisystem anomalies

**DOI:** 10.1101/2024.07.18.24310513

**Authors:** He Fang, Stephen M. Eacker, Yu Wu, Whitney Neufeld-Kaiser, Mercy Laurino, Siobán Keel, Marshall S. Horwitz, Yajuan J. Liu

**Affiliations:** Department of Laboratory Medicine & Pathology, University of Washington, Seattle, WA 98195; Phase Genomics, Seattle, WA 98109; Department of Genetics & Prevention, Fred Hutchinson Cancer Center, Seattle, WA 98109; Department of Medicine, Division of Hematology, University of Washington, Seattle, WA 98112

**Keywords:** complex chromosomal rearrangements, Hi-C, genomic proximity mapping (GPM), optical genome mapping (OGM)

## Abstract

**Purpose:** Complex chromosomal rearrangements (CCRs) are rare structural variants involving three or more chromosomal breakpoints. Most de novo reported CCRs pose challenges for diagnosis and management. They often require karyotyping, fluorescence in situ hybridization (FISH), and chromosomal microarray analysis (CMA) for clinical diagnosis because of the limitations of each method. Here we report an inherited exceptionally complex CCR involving 4 chromosomes and 11 breakpoints in a family with multisystem anomalies.

**Methods:** We evaluated the CCRs using karyotyping, FISH, CMA, and two emerging genomic technologies: high-throughput chromosome conformation capture sequencing (Hi-C; aka genomic proximity mapping, GPM) and optical genome mapping (OGM). We also performed functional studies using transcriptome and methylome analyses.

**Results:** The proband, who had intellectual disability and immune deficiency, shared CCRs with her unaffected mother involving chromosomes 1, 7, and 11 by karyotyping. However, CMA revealed a duplication and three deletions in the proband in contrast to her mother’s balanced genome. Hi-C (GPM) and OGM detected the CCRs and copy number alterations but also uncovered additional breakpoints at high resolution, including an insertion in 4p and two cryptic rearrangements at 7p. Transcriptome and methylome analyses identified likely biological pathways associated with the proband’s phenotypes.

**Conclusion:** Combining cytogenetic and genomic methods provided comprehensive characterization and defined the breakpoints at high resolution in both proband and mother. This underscores the value of novel cytogenetic and genomic techniques in deciphering complex genome rearrangements and the significance of integrative genomic analysis and functional characterization in understanding clinical phenotypes.

## INTRODUCTION

Constitutional complex chromosomal rearrangements (CCRs) usually involve at least two chromosomes and three breakpoints resulting in exchanges of chromosomal segments. CCRs involving 4 chromosomes with 5 breakpoints are classified as exceptional and can be highly complex(1). Phenotypes of people with CCRs vary and the likelihood of a phenotypic abnormality increases with the number of breakpoints involved in apparently balanced CCRs(2, 3). Abnormal phenotypes from CCRs can result from disruption of dosage-sensitive genes, cryptic genomic imbalances near the breakpoints, alteration of the expression of disease-candidate genes, and unmasking of recessive variants on the intact chromosome(4–6). Most familial transmission of CCRs is through the mother(7, 8).

Precise characterization of CCRs is crucial. Identifying disrupted genes or regulatory elements allows for better patient care and genetic counseling and deepens our understanding of CCR etiology. Conventional cytogenetic tools all have specific limitations in investigating CCRs. Karyotyping is a single cell whole genome assay with limited resolution; fluorescence in situ hybridization (FISH) is a targeted assay with limited coverage; and chromosomal single nucleotide polymorphism (SNP) microarray analysis (CMA) can detect copy number variants (CNVs) and large copy-neural regions of homozygosity with high resolution but cannot detect balanced rearrangements(9–11).

High-throughput chromosome conformation capture sequencing (Hi-C; aka genomic proximity mapping, GPM) and Optical genome mapping (OGM) are two technologies that capture ultra-long-range contiguity information and detect all types of structural variants (SVs) in a single assay(12). OGM is an imaging-based method that produces DNA fingerprints spanning very large genomic regions(13). Hi-C (GPM) is a chromatin conformation analysis that identifies chromatin contacts within the nucleus by proximity ligation followed by next-generation sequencing (NGS)(14). We show here that Hi-C (GPM) and OGM, though based on different principles, are both powerful tools to precisely define the breakpoints of inherited CCRs involving chromosomes 1, 4, 7, and 11 in this family.

CCRs may exert a pathogenic effect by gene dosage-dependent mechanisms or through disruption of the genomic architecture that predominantly affect gene expression(15, 16). Such inferred mechanisms of pathogenicity need corroboration by mRNA sequencing. CCRs can also reshape epigenetic landscapes including DNA folding, methylation, and post-translational modifications leading to disease. We hypothesized that investigating transcriptomic and epigenetic landscapes could explain the pathogenic consequences of the CCRs in this family.

## MATERIALS AND METHODS

### A. Proband Consents

Written informed consent was obtained from the proband and the proband’s mother. They were enrolled in this study under a protocol approved by the Institutional Review Board of the University of Washington. Peripheral blood samples were obtained for DNA and RNA studies.

### B. Cytogenetics and Chromosomal SNP Microarray Analysis (CMA)

Karyotyping was performed on phytohemagglutinin (PHA) stimulated peripheral blood lymphocytes according to standard procedures. Twenty GTG-banded metaphases were analyzed. Metaphase FISH analysis was performed on cultured peripheral blood cells using subtelomere probes for chromosomes 1p, 1q, 7p, 7q, 11p, and 11q. CMA of genomic DNA prepared from peripheral blood was performed using the Illumina Infinium CytoSNP-850K BeadChip v1.1. Microarray data were visualized and analyzed using Illumina BlueFuse Multi v4.4 (Illumina Inc., USA) and NxClinical version 10.0 (Biodiscovery Inc., USA).

### C. High-throughput chromosome conformation capture sequencing (Hi-C; genomic proximity mapping GPM)

Hi-C (GPM) libraries were generated using the Phase Genomics Proximo Human kit v4.0 (Phase Genomics Inc., USA) following the manufacturer’s protocol. In brief, white cell pellets from the peripheral blood samples were crosslinked preserving the chromatin structure within the intact nucleus. Following cell lysis, chromatin was immobilized on magnetic beads and digested using restriction enzymes. The overhangs were filled in with biotinylated nucleotides and subjected to proximity ligation. Ligated junctions were purified using streptavidin beads and converted to a standard dual-indexed Illumina-compatible library. In total, we sequenced 200 million Hi-C reads for each of the proband and her mother. Paired-end sequencing data were processed using the Juicer pipeline. The frequency of spatial contact is represented with heatmaps. Hi-C matrices were corrected afterward using “hic_CorrectMatrix” tools from HiCExplorer v1.8.1. Topologically Associating Domain (TAD) boundaries were called using the “hicFindTADs” tool from HiCEx-plorer v1.8.1. First eigenvector (PC1) corresponding to active (A) and inactive (B) compartments was computed using “hicPCA -noe1 –norm” from HiCExplorer after removal of heterochromatic chromosome ends. The correct orientation of PC1, that is, positive values corresponding to the active compartment (A) and negative values corresponding to the inactive compartment (B), was verified for each chromosome using publicly available ENCODE ChIP-seq data.

### D. Optical Genome Mapping (OGM)

Ultra-high molecular weight (UHMW) DNA was extracted from white blood cells and labeled following the manufacturer’s protocols (Bionano Genomics, USA). The fluorescently labeled DNA molecules were loaded on flowcells and imaged sequentially across nanochannels on a Saphyr instrument. A median coverage of > 100x was achieved for both samples. The proprietary OGM-specific software – Bionano Access and Solve (versions 1.6/1.7 and 3.6/3.7 respectively) were used for data processing. De novo assembly was performed using Bionano’s custom assembler software program based on the Overlap-Layout-Consensus paradigm. SVs were identified based on the alignment profiles between the de novo assembled genome maps and the Human Genome Reference Consortium GRCh38 assembly. Fractional copy number analyses were performed from alignment of molecules and labels against GRCh38.

### E. mRNA-Seq

Library preparation for RNA-seq was performed on 100 ng of total RNA isolated from peripheral blood using TruSeq® Stranded mRNA Sample preparation kit (Illumina Inc., USA). The library’s size distribution was validated and quality inspected on a Bioanalyzer TapeStation (Agilent Technologies). High quality libraries are pooled for 75 bp paired-end sequencing on a NextSeq500 instrument (2 × 75 cycles) according to manufacturer instructions (Illumina Inc., USA).

Reads were aligned to the human annotation reference genome GRCh38 using STAR 2.5.2b. BAM files of mapped reads were visualized by the integrative genomics viewer (IGV). A total of 62 and 71 million (M) uniquely mapped reads (mappability: > 84%) were obtained for the proband and her mother respectively. RNA-seq data was compared to public mRNA-seq data sets from whole blood of 6 females age matched to the proband and her mother (RNA-seq whole blood of Dutch 500FG cohort, National Library of Medicine National Center for Biotechnology Information Gene Expression Omnibus GSE134080). Libraries for the external controls were prepared using TruSeq mRNA Sample prep kit. The single-end read-length is 100 bp. These control data sets have a similar sequence depth and quality with 25 million uniquely mapped reads (mappability: > 87%) which were reprocessed with our own data for normalization and comparison.

Differential expression profile analyses were done with weighted trimmed mean of M-values (TMM) normalization method and GENCODE gene annotation in the EdgeR statistical software package (Bioconductor) to investigate the relative change in gene expression (i.e., normalized counts) between different samples. Two differential expression (DE) comparisons were conducted: (1) the proband versus three healthy age-matched female controls; (2) proband’s mother versus three healthy age-matched female controls. 11460 expressed genes with a median of over five raw counts per million (CPM) for the eight samples (the proband, proband’s mother, and six control females) were included for DE analyses. Absolute expression fold changes of two and false discovery rate (FDR) < 0.01 were set as the threshold to call genes with significantly differentially expressed genes (DEGs; Supplementary Table 1). DEGs were used for gene ontology (GO) enrichment analysis.

Gene ontology overrepresentation test was done by PANTHER to test whether upregulated or downregulated DE genes are enriched in GO terms (e.g., biological processes and pathways) compared to the reference of 20996 human genes. FDR < 0.05 from Fisher’s Exact test was used for the cutoff. Biological processes or pathways with enrichment fold less than one were not shown.

### F. Methylation array data processing

We performed methylome analysis with the proband’s peripheral blood samples and an age matched normal female control using Illumina Infinium Human Methylation EPIC BeadChip (Illumina Inc., USA). This methylation array covers over 850000 methylation sites across the genome at single-nucleotide resolution. DNA methylation profiling was performed according to the manufacturer’s instructions. We performed differential DNA methylation analysis using a customized R package pipeline. We stratified quantile normalized data using the ‘minfi’ ‘preprocessQuantile’ function. After that, probes targeting sex chromosomes, containing single-nucleotide polymorphisms, not uniquely matching, as well as known crossreactive probes, were removed. Most significantly differentially methylated CpG were identified by fitting a regression model with the disease as the target variable using the ‘limma’ R package. Differentially methylated gene lists identified through methylation array analysis were used for gene ontology (GO) enrichment analysis. Gene ontology overrepresentation test was done by PANTHER to test whether up-regulated or downregulated DE genes are enriched in GO terms (e.g., biological processes and pathways) compared to the reference of 20996 human genes. FDR < 0.05 from Fisher’s Exact test was used for the cutoff. Biological processes or pathways with enrichment fold less than one are not shown.

## RESULTS

### G. Clinical presentation

The proband is a 30-40 year old female with a history of complex immunological disorders and developmental delay.

The proband was the product of a term pregnancy. Maternal exposures were limited to low dose aspirin prescribed for recurrent pregnancy loss. The proband was diagnosed in childhood with developmental delay without a specific recognizable syndrome based on Wechsler Intelligence Scale testing. The proband completed high school through a special education program, remains independent in activities of daily living, reads, watches television, and lives with others in an adult family home.

Physical examination revealed a pleasant, appropriately verbally communicative female. The proband exhibited subtle midface hypoplasia with epicanthic folds, hypertelorism, an appearance of a webbed neck, and triangular shaped fingers. Proband’s mother is a healthy, 60-70 year old female with a history of 2 miscarriages with a partner different from the proband’s father. She completed high school, has been professionally employed, and has an unremarkable medical history with no known autoimmune or other disorders and an unremarkable physical examination.

### H. Cytogenetics and Chromosomal SNP Microarray Analysis (CMA) Results

The proband’s karyotype analysis revealed a complex but seemingly balanced set of rearrangements: 46XXder(1)t(1;7)(q42.2;p22)der(7)t(1;7)(q44;q11.2)der(11) (pter->11q25::1q42->1q44::7q11.2->7qter) (Figure 1A). This exceptionally complex chromosomal rearrangement involves translocations of chromosomes 1, 7, and 11 with breakpoints at 1q42.2, 1q44, 7p22, 7q11.2, 11q25 respectively.

**Fig. 1.**
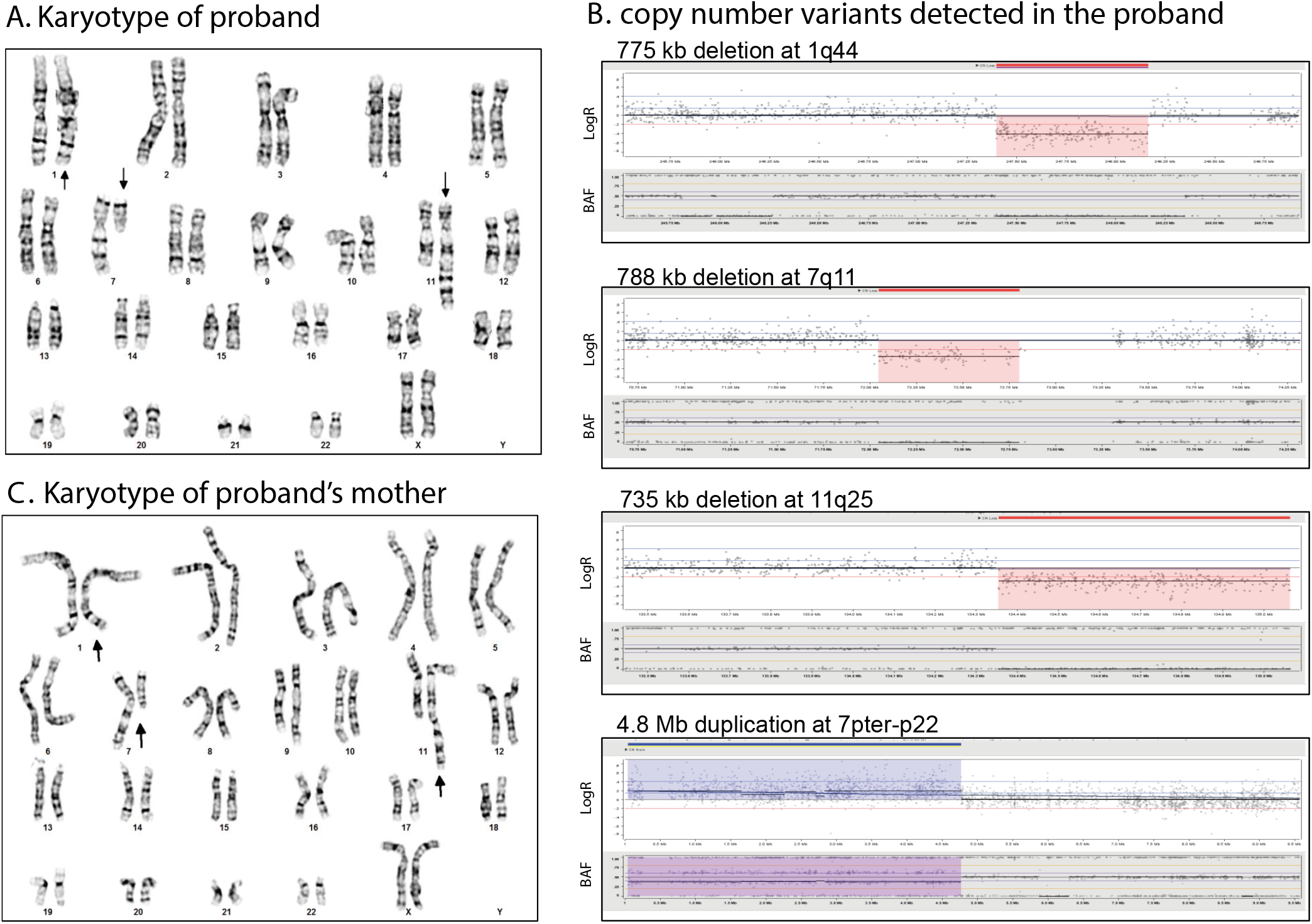
Results of G-banded chromosome analysis and SNP genomic microarray analysis of the proband and the proband’s mother. (A) Karyotype of the proband. (B) Karyotype of the proband’s mother. (C) Four copy number variants detected in the proband by SNP genomic microarray analysis with genomic coordinates on the X axis and Log2R and BAF on the Y axis (see Table 1 for details)

To determine whether the CCR was truly balanced, we performed genome-wide CMA. This revealed four copy number variants (CNV): a 4.8 Mb terminal duplication of 7pterp22 and three deletions of 774 kb, 788 kb, and 675 kb from 1q44, 7q11, and 11q25-qter respectively (Figure 1B, Table 1). None of the CNVs were inherently pathogenic. The 7q11 deletion does not overlap the Williams syndrome critical region; the 11q25-qter deletion does not contain the critical region for Jacobsen syndrome, and the 1q44 deletion does not encompass the critical region for 1q43q44 microdeletion syndrome.

**Table 1.** Copy number variations detected by chromosomal SNP microarray analysis (CMA), Hi-C, and OGM in the proband.

| [hg38] | Loss on Chr1 | Loss on Chr7 | Gain on Chr7 | Loss on Chr11 |
| --- | --- | --- | --- | --- |
| **CMA** | chr1:247396871-248171504 | chr7:1-4799433 | chr7:72046323-72834567 | chr11:134351449-135086622 |
| **Hi-C** | chr1:247415001-248170001 | chr7:1-4785001 | chr7:72055001-72840001 | chr11:134350001-135070000 |
| **OGM** | chr1:247286043-248164170 | chr7:706747-4738703 | chr7:71995945-72863337 | chr11:134309117-135069566 |

The detection of CNVs in the proband showed that the proband’s CCR was actually unbalanced. The terminal duplication at 7pter-p22 likely encompasses the breakpoints on the derivative chromosome 1 as identified by karyotype. Similarly, the deletions at 1q44, 7q11, and 11q25-qter likely encompass the breakpoints on the derivative chromosomes 7 and 11. To determine if the proband’s CCR was inherited or de novo, a peripheral blood sample from the proband’s mother was requested for karyotyping and CMA. The proband’s father was not available for testing. Surprisingly, the proband mother’s karyotype seemed indistinguishable from the proband with complex rearrangements involving chromosome 1, 7, and 11 (Figure 1C). In contrast, the proband’s mother’s CMA revealed no detectable copy number variants (Supplementary Figure 1). This finding is consistent with the mother’s unremarkable clinical presentation and suggested that cryptic rearrangements were present which fall beyond the detection limits of karyotyping (3-10 Mb resolution), particularly in subtelomeric chromosomal regions. Metaphase FISH was performed using subtelomeric probes that hybridize to 1pter, 1qter, 7pter, 7qter, 11pter, and 11qter (Figure 2). Consistent with the proband’s mother’s normal CMA results, two signals each were seen for 1qter on chromosome 1 and derivative chromosome 7, 7pter on chromosome 7 and derivative chromosome 1, and 11qter on chromosome 11 and derivative chromosome 7 (Figure 2A). In contrast, the proband had two signals of 1qter on chromosome 1 and derivative chromosome 7 but three signals of 7pter on chromosome 7, derivative chromosome 7, and derivative chromosome 1, and one signal of 11qter on chromosome 11 (not on derivative chromosome 7 as in her mother) (Figure 2B). The FISH results confirmed the unbalanced nature of the CCR in the proband and was consistent with the CMA results.

**Fig. 2.**
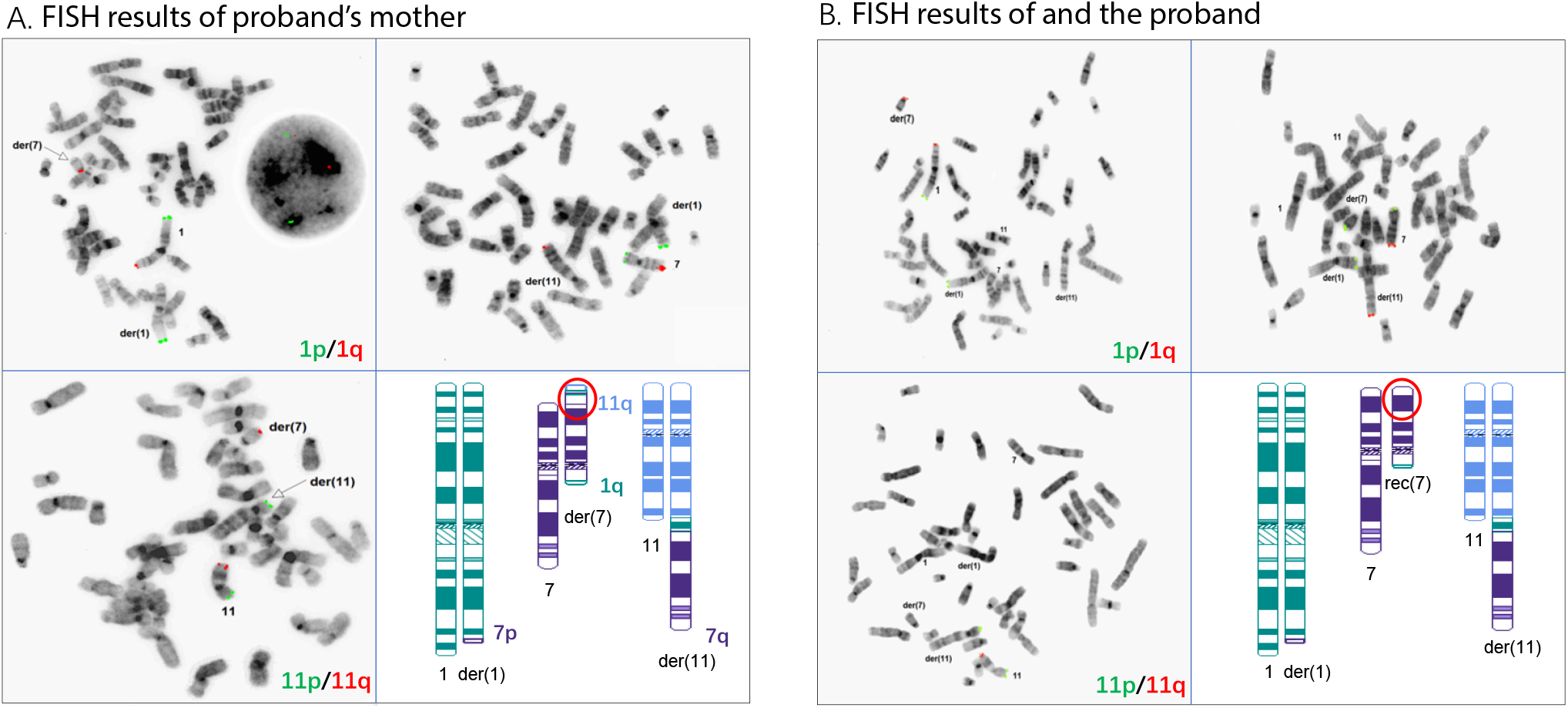
FISH analysis results of the proband’s mother (A) and the proband (B). Due color FISH using sub-telomeric probes set on metaphase cells identified localizations of 1p (green) and 1q (red), 7p (green) and 7q (red), 11p (green) and 11q (red) on chromosome 1, the der(1) chromosome 7, the der(7), chromosome 11, and the der(11). Schematic diagrams in the lower right corner of (A) and (B) show localization of FISH signals on ideograms of the respective rearranged chromosomes.

### I. CCR Breakpoint Mapping by Hi-C (GPM) and OGM

Hi-C (GPM) and OGM were performed to precisely characterize the breakpoints of both unbalanced and balanced CCRs in the proband and her mother. OGM simultaneously identified all four CNVs (Table 1) and the structural variants involving chromosomes 1, 7, and 11 in the proband (Figure 3A). The breakpoints detected by OGM were 1q42, 1q44, 7p22, 7q11.2, and 11q25, consistent with the karyotype (Figure 3B). OGM also clarified the precise breakpoints and genes disrupted in the proband including ARID4B, GLB1L2, URB2, and AP5Z1 (Table 2). These rearrangements correspond to the aberrations we observed on der(1), der(7), and der(7) via karyotype. Moreover, a novel balanced insertion between chromosome 4 and chromosome 7, which was not observed by any conventional cytogenetic test method, was identified (Figure 3A, 3B).

**Table 2.** Breakpoint coordinates detected by OGM and Hi-C in the proband and proband’s mother.

|  | Translocations detected by OGM [hg38] |  | Translocations detected by Hi-C [hg38] |  |
| --- | --- | --- | --- | --- |
|  | chrA | chrB | chrA | chrB |
| **Proband** |  |  |  |  |
| t(1;11) | chr1:235309987 | chr11:134339794 | chr1:235305001 | chr11:134380001 |
| t(4;7) | chr4:7484910 | chr7:71350922 | chr4:7484910 | chr7:71335001 |
| t(4;7) | chr4:7498374 | chr7:71330734 | chr4:7484910 | chr7:71335001 |
| t(1;7) | chr1:229627661 | chr7:4784641.5 | chr1:229625001 | chr7:4780001 |
| t(1;7) | chr1:248174298 | chr7:67453424 | chr1:248170001 | chr7:67460001 |
| **Proband's mother** |  |  |  |  |
| t(1;11) | chr1:235309987 | chr11:134339794 | chr1:235305001 | chr11:134380001 |
| t(1;11) | chr1:247397537 | chr11:134350029 | chr1:247390001 | chr11:134345001 |
| t(4;7) | chr4:7484910 | chr7:71350922 | chr4:7485001 | chr7:71351001 |
| t(4;7) | chr4:7498374 | chr7:71321506 | chr4:7498401 | chr7:71321501 |
| t(1;7) | chr1:229627661 | chr7:4784641.5 | chr1:229625001 | chr7:4780001 |
| t(1;7) | chr1:248168445 | chr7:6121028.5 | chr1:248170001 | chr7:6085001 |
| t(1;7) | chr1:248191628.5 | chr7:67453424 | chr1:248170001 | chr7:67460001 |
| t(7;7) | chr7:72067101 | chr7:6085026 | chr7:72840001 | chr7:6080000 |
| t(7;7) | chr7:72841562 | chr7:6483388 | chr7:72840001 | chr7:6350000 |

**Fig. 3.**
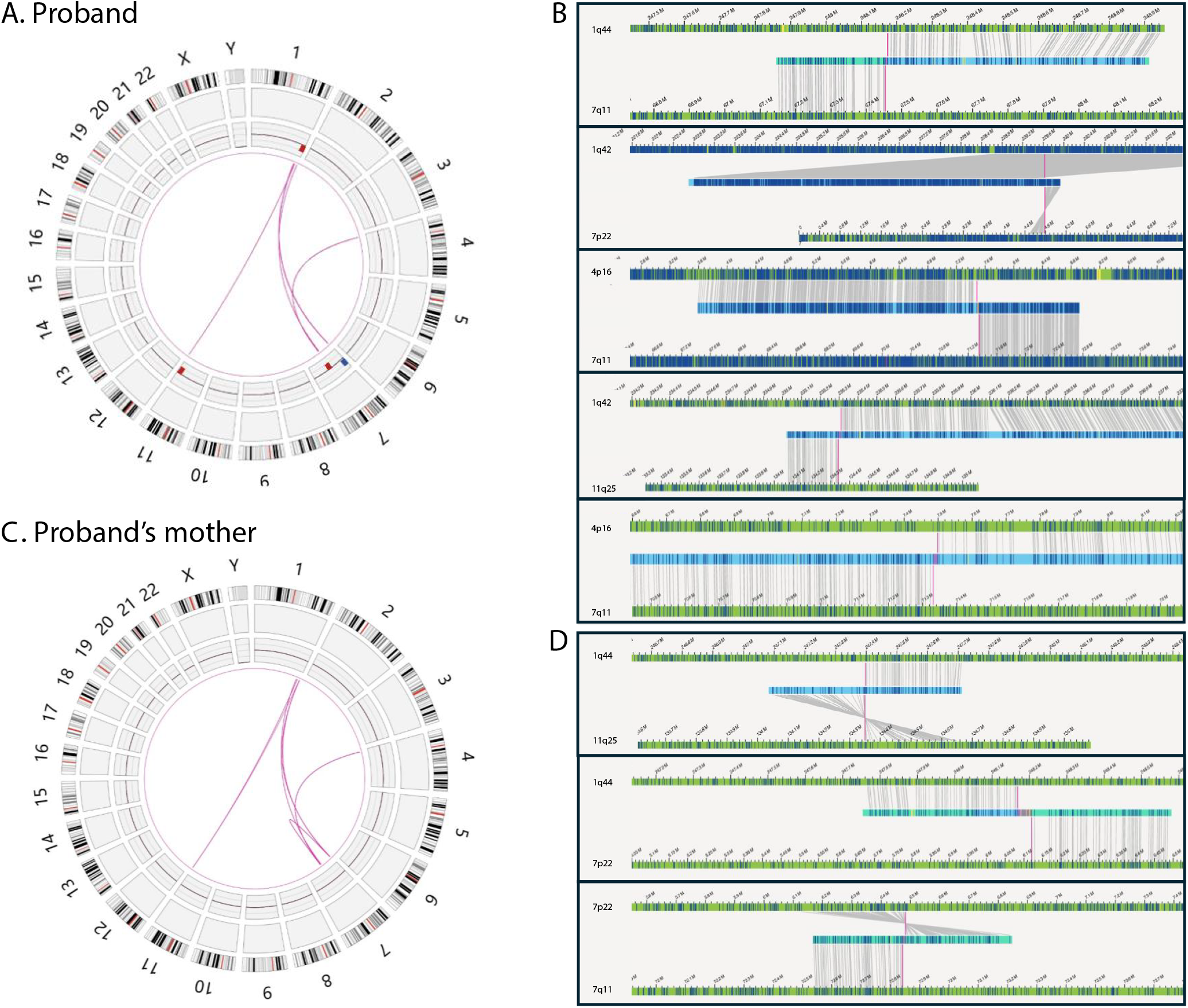
OGM results in the proband and her mother. Circos plots display the CCRs in the proband (A) and the proband’s mother (C). The outer rings represent chromosomes 1-22, X, and Y; inner rings show the CCRs detected. Circos tracks include: Cytoband, SV track, CNV track (red-deletion, blue-duplication), and translocations. Detailed views of rearrangements around the breakpoints (B, D). In the proband, a total of 5 translocations were shown (B). In the proband’s mother, 8 translocations observed including 5 same as those in the proband (C) and 3 in the proband’s mother only (D).

No CNVs were identified in the proband’s mother by OGM, consistent with the findings by CMA. The translocation events in the proband’s mother are more complicated. Nine rearrangements were identified in total, including 7 interchromosomal rearrangements and 2 intrachromosomal rearrangements (Figure 3C). All 5 interchromosomal rearrangements found in the proband are also present in her mother, but the latter has two additional rearrangements between 1q44 and 11q25, and 1q44 and 7p22 (Figure 3D). The intrachromosomal rearrangements were only found in the proband’s mother and formed within chromosome 7 (between 7p22 and 7q11) (Figure 3D). These results suggest that the majority of the differences between the proband and her mother are associated with 1q44, 11q25, and 7p22, supporting our previous FISH findings.

The high-resolution Hi-C (GPM) assay verified the proband’s CNVs as identified by CMA and OGM (Table 1). Hi-C (GPM) also successfully detected the translocations between chromosome 1, 7, and 11 and the novel insertion between chromosome 4 and chromosome 7 at a high resolution (Figure 4A). In the proband, all 5 rearrangements were detected by Hi-C (GPM) (Figure 4B), and the coordinates are listed in Table 2. To provide a better illustration, each translocation revealed by the heatmap is presented by an ideogram. For example, we observed abnormal stripes (as indicated by a, b, and c) on the contact map between chromosome 1 and chromosome 7 (Figure 4B, top left). The stripe indicated by an arrow shows an inter-chromosomal interaction between chromosome 7 (0 Mb-8 Mb) and chromosome 1 (0 Mb-235 Mb), suggesting there is a translocation of 7p22 to 1q42. Similarly, from stripes b and c, we can infer two additional rearrangements between fragments of chromosome 1 and chromosome 7. Of note, only the rearrangements represented by the relevant chromosomes are taken into consideration to construct the ideogram up until this point of the analysis. Overall, these rearrangements are concordant with the aberrations we observed by OGM.

**Fig. 4.**
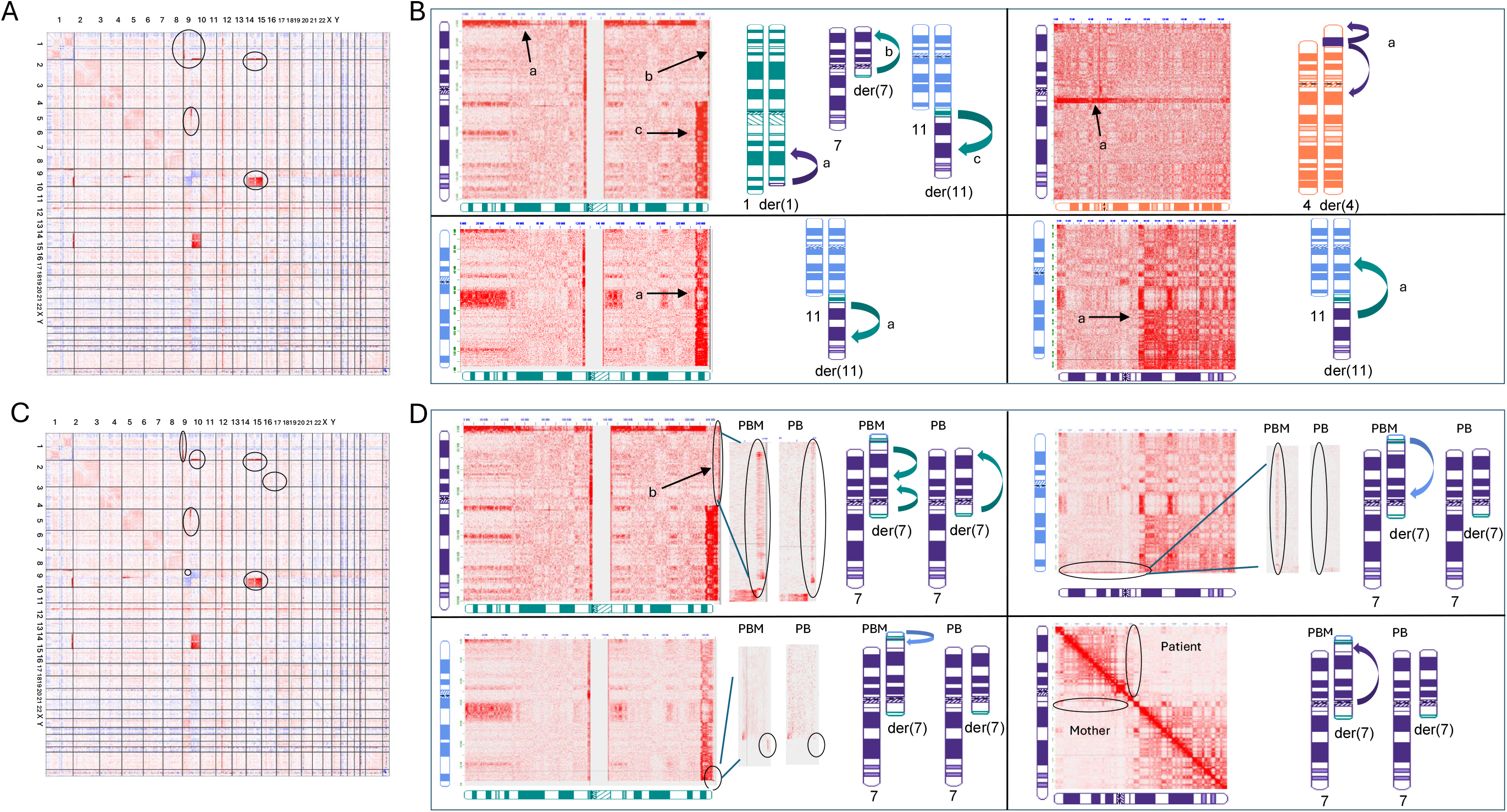
Hi-C (GPM) results in the proband (A, B) and her mother (C, D). (A, C) Genome-wide differential Hi-C heatmaps represent the CCRs in the proband and her mother when compared to a normal female control. Red color indicates gain of interactions in the sample and blue color indicates loss of interactions in the sample. Chromosome by chromosome view of Hi-C (heatmaps in the proband (B) and her mother (D): four pairs of inter-chromosomal interactions were shown for the proband (B) including chromosome 1 with 7, chromosome 1 with 11, chromosome 4 with 7, and chromosome 7 with 11; three pairs of inter-chromosomal interactions and one pair of intra-chromosomal interactions specifically observed in the proband’s mother (D). Each left panel in (B) and (D) shows the original heatmap and the right panel shows the representative ideogram of the translocation.

We also examined the CCR in the proband’s mother by Hi-C (GPM). No CNVs were identified, consistent with the findings by OGM and CMA. In addition, the genome-wide Hi-C (GPM) interaction heatmap looks very similar to the proband, showing all the rearrangements observed in the proband (Figure 4C). The subtle differences in the proband’s mother’s specific rearrangements became more evident when we examined the contact heatmap chromosome by chromosome (Figure 4D). For example, we have examined the translocation between chromosome 1q44 to chromosome 7q22 by stripe b in the proband. When we zoomed in, we observed clear differences in the contact pattern in the proband’s mother as shown in Figure 4D. The stripe is composed of 3 components here, including a stripe between 1q44 (248 Mb-250 Mb) and chromosome 7 (67 Mb-5 Mb), a stripe between 1q44 (247 Mb-248 Mb) to chromosome 7 (5 Mb-67 Mb, different orientation), and a small block between 1q44 (247 Mb-250 Mb) and chromosome 7 (72 Mb-73 Mb). With this information, we can infer complicated translocations as shown in the ideogram in Figure 4D. The additional translocations identified in the proband’s mother by OGM, including interactions between 1q44 and 7p22, 1q44 and 11q25, and 7p22 and 7q11, were also evident by Hi-C (GPM) (Figure 4D).

The long-range interactions between the translocated segments and distant neighboring genomic regions detected by Hi-C (GPM) in addition to the interactions directly generated by the conjugated segments provide us with supporting information related to the chromosomal rearrangement structure. For example, the proband’s mother had two stripes of contacts between 1q44 and chromosome 7. One of the stripes showed a darker signal toward 7p, indicating that this fragment is translocated to 7p. The other one showed a darker signal toward 7cen, indicating that this fragment is translocated closer to 7cen. With these considerations, we were able to reconstruct the CCRs in the proband and her mother (Ideogram, Figure 5A, B). Note, both OGM and Hi-C (GPM) are methods to measure genomic segments and they are not capable of mapping the whole genome continuously and directly. Due to the complexity of these rearrangements and the limitation of the detection methods, we cannot rule out other possible rearrangements.

**Fig. 5.**
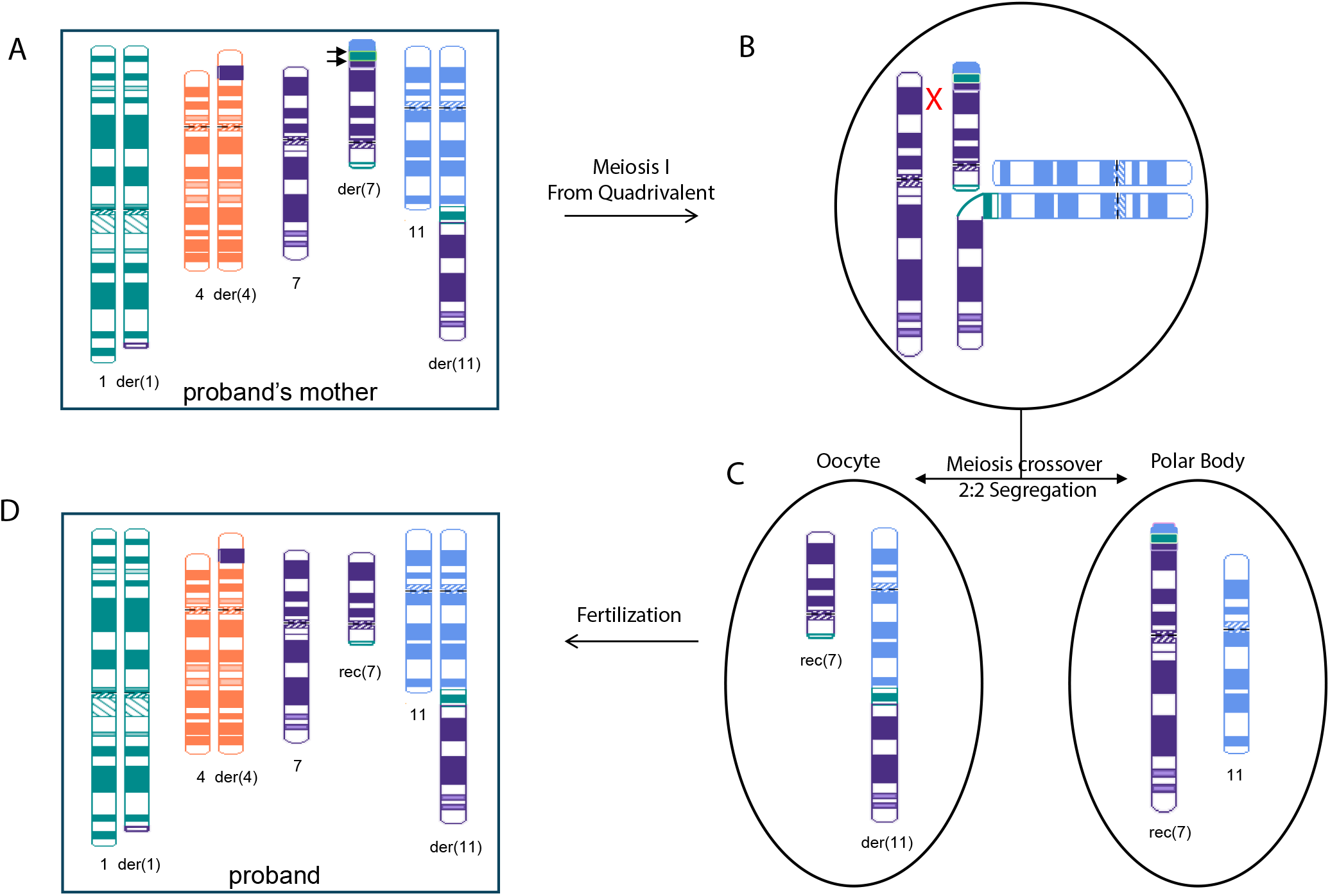
Proposed model of CCR transmission from proband’s mother to the proband resulting in unbalanced chromosomal regions. Postulated CCRs in the proband’s mother (A) and the proband (D) include chromosome 1, 4, 7, and 11. While derivative chromosome 7p in the proband’s mother (A) comprises segments of 1q, 11q, and 7q, recombinant chromosome 7 rec(7) in the proband contains intact 7p sequence. Hypothesized meiotic recombination event in the proband’s mother where three-way CCR heterozygote forms a quadrivalent structure (B). Crossover occurs between homologous 7p and derivative 7p and followed by 2:2 alternative segregation (C).

Overall, we were able to reconstruct the complicated CCRs for both the proband and her mother. We also confirmed that both OGM and Hi-C (GPM) can facilitate robust detection of cryptic balanced and unbalanced translocations in clinical practice with a high degree of accuracy.

### J. Effect of the CCR on Gene Expression

To identify dysregulated genes and pathways associated with the proband’s clinical presentation, we performed transcriptome analysis by mRNA-seq of the proband’s and her mother’s peripheral blood samples. We also compared expression profiles in the proband and her mother with public mRNA-seq data sets from peripheral blood samples of age-matched control females.

We first examined the expression of genes spanning the breakpoints of the CCRs in the proband, including ARID4B, GLB1L2, SORCS2, URB2, and AP5Z1. RNA-seq revealed a hybrid transcription product of ARID4B and GLB1L2, confirming the t(1;11) translocation revealed by other methods (Supplementary Figure 2). The same transcription product was found in the proband’s mother with a similar level of expression and is absent in healthy controls. SORCS2, URB2, and AP5Z1 all have a median expression less than 1 TPM in the proband, her mother, and in healthy control data sets from GSE and the GTEX whole blood database, indicating they are not expressed in whole blood (Supplementary Table 1). We next examined the expression of genes fully contained within the CNVs. There are 156 such genes, including 71 coding genes, 72 noncoding genes, and 13 pseudo genes. Of the 71 coding genes, 23 are in the olfactory gene family (OR genes). None of these 156 genes shows dosage sensitivity according to Clingen (Supplementary Table 2). Differential gene expression analysis showed that only TRIM58 (1q44) varies significantly in the proband compared to her mother and the public data sets (Supplementary Figure 3A). The proband’s TRIM58 expression is 3.15-fold lower than her mother’s and 2.68-fold lower than normal female controls, both with FDR<0.01. The TRIM58 gene is exclusively expressed in late-stage erythroblasts and involved in erythrocyte development. Taken together, these results suggest that most genes spanning the CCR breakpoints or within the CNVs are unlikely to have caused the proband’s phenotype.

To address the effect of the CCR on global gene expression, we identified DEGs (differentially expressed genes) in the proband compared with her mother and age-matched controls. We identified 510 and 757 genes that are up-and down-regulated, respectively (Supplementary Table 3). These genes are uniformly located across chromosomes without significant enrichment on any specific chromosome (Supplementary Figure 3B). Gene ontology (GO) analysis was performed for up-regulated and down-regulated DEGs separately, which has been shown to be more powerful than using all DEGs together to identify disease-associated pathways (Figure 6A). Upregulated DEGs are highly enriched in immune response processes, including Fc receptor mediated inhibitory signaling pathway, interferon-gamma-mediated signaling pathway, and cellular response to type I interferon pathway. These pathways could be associated with the proband’s autoimmune disorders, including systemic lupus erythematosus with Sjogren syndrome, suspected lupus nephritis, and immune thrombocytopenic purpura. In contrast, downregulated genes are enriched in general biological processes, including base-excision repair and mitochondrial RNA metabolic process.

**Fig. 6.**
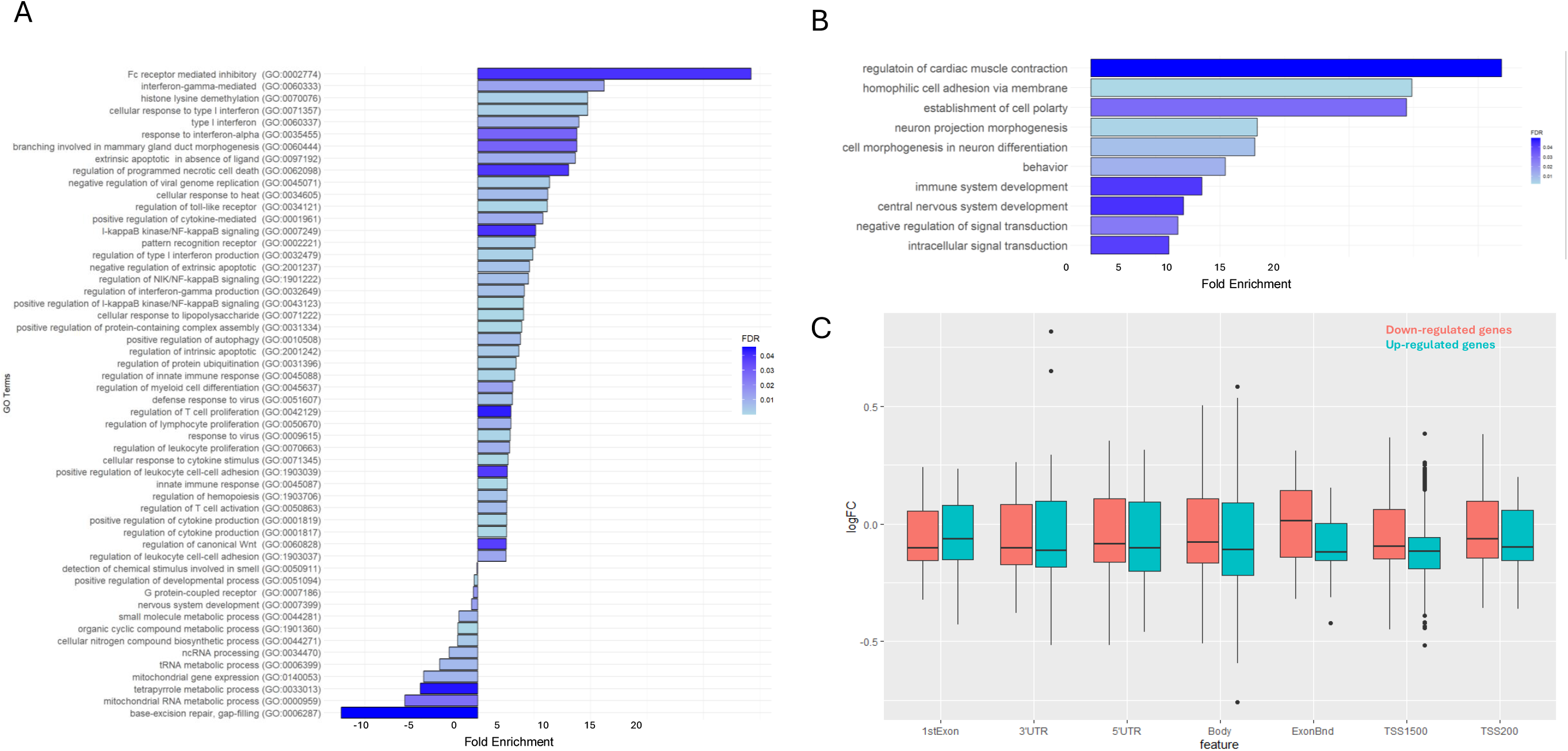
Impact of the CCRs on Gene Expression and DNA methylation. Differential expression analysis of genes in CNV regions by comparing the proband to normal female controls and proband’s mother (orange and blue dots respectively). (A) RNA-seq identified pathways affected in the proband; up-regulated genes in immune response, down-regulated genes in neuronal development. (B) Functional pathways affected identified by top genes enriched by significant CpGs. (C) Correlation of the DNA methylation profile with the RNA expression profile. Red color bar indicates the methylation change of down-regulated genes and blue color bar indicates the methylation change of up-regulated genes.

### K. Effect of the CCR on the Epigenetic Landscape

The epigenetic landscape encompasses multiple key features, including methylation profiles and chromosomal organization, which together contribute to the regulation of gene expression and cellular identity. We first analyzed the methylation profiles of the proband and her mother. The top 5000 CpGs with significantly differential methylation status in the proband were identified, and these CpGs are distributed evenly across the genome (Supplementary Figure 3C). The chromosomes involved in the CCRs (chromosomes 1, 4, 7, and 11) are not enriched for differential CpGs, suggesting the CCR does not lead to major alterations in the proband’s methylation landscape. GO analysis was performed for the top 70 genes associated with these CpG sites (Figure 6B) and showed that the proband’s top genes with differential methylation status are highly enriched in neuronal developmental processes relevant to the proband’s developmental delay, including neuron projection morphogenesis, neuron differentiation, and establishment of cell polarity.

We further correlated the DNA methylation profile with the RNA expression profile. The median methylation level is increased for the TSS (transcriptional start site) regions of downregulated genes and decreased for the upregulated genes, consistent with hypermethylation of downregulated genes and hypomethylation for upregulated genes (Figure 6C). On the other hand, the proband’s differential methylation analysis identified 2142 hypomethylated probes and 235 hypermethylated probes, corresponding to the hypomethylation of TSS regions of 1239 genes and hypermethylation of TSS regions of 126 genes. We did not observe significant change in expression of these genes likely because most of these genes are not expressed in blood cells.

Chromosomal organization is another key aspect of the epigenetic landscape. We first assessed the changes of the proband’s overall chromosomal organization. To do this, we computed the similarities between the genome-wide Hi-C contact matrix of the proband, her mother, and a healthy control with HiCRep, a tool that can provide a statistical evaluation of the Hi-C interaction matrix (Figure Supplementary 4).

The HiC contact matrix in the proband showed high correlations (>0.89) with both the healthy controls and the proband’s mother. Chromosomes involved in the CCR (chromosomes 1, 4, 7, and 11) present with similar correlation compared to other chromosomes, suggesting the translocations do not lead to major alterations in the proband’s chromosomal organizations.

Having established the overall correlation between the Hi-C interaction matrices of the two samples, we next conducted a detailed analysis of the topologically associating domains (TADs) and A/B compartments to investigate the similarities and differences in chromatin organization between the samples. TADs are regions of the genome with high intra-domain chromatin interactions and low inter-domain interactions, thought to play a role in gene regulation and chromatin organization. A/B compartments refer to large-scale chromatin domains that have distinct epigenetic and transcriptional properties, with A compartments enriched for active chromatin marks and highly transcribed genes, and B compartments enriched for repressive marks and lowly expressed genes. We examined the A/B compartments and TAD organizations. No A/B compartment change is observed except the regions around translocation breakpoints (supplementary 4B). The TAD organizations are also maintained in the context of complicated translocations in both the proband and proband’s mother (Figure supplementary 4C).

## DISCUSSION

The frequency of pathogenic CNVs is higher in individuals with developmental delay(17, 18). CMA can efficiently identify CNVs and has become the first-tier test for people with unexplained developmental delay. For example, a study of 329 people with intellectual disability confirms that causative CNVs are frequently found even in cases of mild intellectual disability(19). The proband in our study is a female with delayed speech, cognitive impairment, and autoimmune disorders. CMA identified a 768 kb deletion of 1q44 that affects NLRP3, GCSAML, and OR, a 788 kb deletion of 7q11.22 to 7q11.23 that affects CALN1 and TYW1B, a 709 kb deletion of 11q25 that affects GLB1L2 and B3GAT1, and a 4.7 Mb duplication of 7p22.3 to 7p22.1 involving 37 genes. The CNVs do not overlap any known dosage-sensitive genes or regions, and the classification of each of these regions is uncertain per ACMGG interpretation standards(20). Thus, these CNVs may be associated with the proband’s phenotype, but more evidence is needed to determine if the chromosomal rearrangements are related to the underlying molecular mechanism of the proband’s condition.

Further investigation revealed the inherited nature of the proband’s CCR by both conventional and novel cytogenetic methods. Conventional karyotype analysis revealed that the proband and her mother had nearly indistinguishable chromosomal abnormalities characterized by three translocations involving three different chromosomes. Additional FISH analysis successfully identified specific chromosomal rearrangements localized to chromosomal 7p in the proband and proband’s mother but did not precisely localize the breakpoints. CMA effectively identified the coordinates of the breakpoints of the imbalanced rearranged chromosomal segments. Still, without further details, the chromosomal rearrangements could not be fully resolved in this family. Both OGM and Hi-C (GPM) identified a novel translocation between chromosomal 4 and chromosomal 7 in the proband and her mother. OGM and Hi-C (GPM) also uncovered a set of unique rearrangements only carried by the proband’s mother, including an intra-chromosomal translocation within chromosome 7, an additional translocation between chromosome 1 and chromosome 11, and an additional translocation between chromosome 1 and chromosome 7. These observations, especially the translocations exclusively detected in the proband’s mother, lead us to suspect a more complex rearrangement exists in the proband’s mother and that such rearrangement spans chromosomes 1, 4, 7, and 11. Moreover, Hi-C (GPM) was able to show relative distances between translocated segments, so we were able to extrapolate that the unique sets of rearrangement events carried by the proband’s mother are sequentially located on the chromosome 7p arm. Overall, we reconstructed the CCR in the proband and her mother. We propose a model to explain how the CCR was passed from the proband’s mother to the proband, leading to multiple unbalanced chromosomal regions (Figure 5). We suspect that there was a recombination event between the normal and derivative chromosomes 7 at 7p during meiosis 1 pairing in the proband’s mother, involving chromosome 7, the derivative chromosome 7, chromosome 11, and the derivative chromosome 11. During meiosis I, the three-way CCR heterozygote came together to form a quadrivalent configuration (Figure 5B). Meiotic crossover occurred between homologous chromosome 7p and derivative chromosome 7p, followed by 2:2 alternative segregation (Figure 5C). Thus, the recombinant chromosome 7 in the proband (Figure 5D) has a normal chromosome 7p, while the der(7) in her mother (Figure 5A) has a 7p composed of part of 1q, 11q, and 7q. This crossover would also explain the CNVs observed in the proband while her mother is balanced.

In addition, we investigated the genetic cause of the proband’s clinical presentations. With OGM and Hi-C (GPM), we identified a total of 8 breakpoints and 4 regions with CNVs in the proband. Interestingly, there are 13 breakpoints in total in the proband’s mother, including the 8 breakpoints in the daughter and an additional 5 breakpoints found only in the proband’s mother with no CNVs. Considering that the proband’s mother is phenotypically unremarkable, we suspect that the breakpoints are unlikely to account for the proband’s phenotype. Instead, the CNVs and their interactions with the global transcriptome and epigenetic landscape are likely to be responsible. We performed transcriptome and methylome analysis in the proband. The expression and methylation profiles of the genes directly located within the CNV or spanning the breakpoints were unaffected in the proband. In contrast, we identified functional pathways potentially associated with the proband’s phenotype using the global differential expression and methylation profiles, suggesting it is the disruption of cellular networks that leads to the proband’s findings. For example, we identified upregulated genes highly enriched in pathways associated with immune response processes, including Fc receptor mediated inhibitory signaling pathway, interferon-gamma-mediated signaling pathway, and cellular response to type I interferon pathway. These pathways could be associated with the proband’s complex immune phenotype. Similarly, the proband’s top genes with differential methylation status are highly enriched in neuronal developmental processes relevant to the proband’s developmental delay, including neuron projection morphogenesis, neuron differentiation, and establishment of cell polarity. In addition, the proband’s transcriptome and methylome profiles correlated with the TSS regions hypermethylated for downregulated genes and hypomethylated for the upregulated genes. While the transcriptome analysis identified a set of genes potentially relevant to the proband’s immune deficiency, the methylome analysis identified a set of genes potentially relevant to the proband’s developmental delay. Although the differentially expressed genes identified by RNA-seq did not overlap with the differentially methylated genes identified by DNA methylation array, it is known that changes in gene expression may not always correlate directly with DNA methylation status, as other regulatory elements can modulate gene expression independently of methylation. Further studies are required to determine the significance of these differences in expression and methylation patterns.

In summary, this study demonstrates the power of advanced cytogenetic methods, including OGM and Hi-C (GPM), in resolving CCRs and their pathophysiologic consequences. Conventional cytogenetic methods are routinely applied in the cytogenetics lab. While these are powerful diagnostic tools, they also have limitations, particularly in defining CCRs with limited resolution, as seen with this case. This study showcases the unique strengths and limitations of each method (Table 3) and provides an example of how the complementary approaches of OGM, high-throughput Hi-C (GPM), and conventional cytogenetics provide a comprehensive characterization of complex structural variants. Our approach also highlights how RNA-seq and methylome analyses can further inform our understanding of the molecular underpinnings of phenotypes of patients with CCRs.

**Table 3.** Strength and Limitation of FISH, Karyotype, chromosomal SNP microarray (CMA), OGM, and Hi-C (GPM) in the detection of copy number variants (CNV) and structural variant (SV) including complex chromosomal rearrangements (CCRs).

|  | FISH | Karyotype | CMA | OGM | Hi-C (GPM) |
| --- | --- | --- | --- | --- | --- |
| **Region** | targeted region | genome-wide | genome-wide | genome-wide | genome-wide |
| **Resolution** | 100 kb (medium) | 5 Mb (low) | 40 kb (high) | 40 kb (high) | 40 kb (high) |
| **Detection of CNV** | Yes | Yes | Yes | Yes | Yes |
| **Unbalanced SV** | Yes | Yes | Yes | Yes | Yes |
| **Balanced SV** | Yes | Yes | No | Yes | Yes |
| **Reconstruction of complex CCR** | Yes | Yes | No | No | Yes |

## Data Availability

All data produced in the present study are available upon reasonable request to the authors.

## Acknowledgements

This work was funded in part by a grant from the Brotman Baty Institute (BBI) to YJL and HF, NICHD/NIH R44HD104323 to SME.

## Author Contributions

Conceptualization: Y.J.L., S.M.E., H.F.; Data analysis and curation: H.F., Y.J.L., S.M.E., Y.W.; Clinical evaluation: M.S.H., S.K., M.L.; Funding acquisition: a Brotman Baty Institute grant to H.F., Y.J.L.; NICHD/NIH R44HD104323 to S.M.E.; Writing-original draft: H.F., Y.J.L., S.M.E.; Writing-review & editing: all.

## Ethics Declaration

This project was approved by the institutional review board (IRB) of the University of Washington. Written informed consent was obtained from all participants or their parents.

